# Seroprevalence of anti-SARS-CoV-2 IgG at the epidemic peak in French Guiana

**DOI:** 10.1101/2020.09.27.20202465

**Authors:** Claude Flamand, Antoine Enfissi, Sarah Bailly, Christelle Alves Sarmento, Emmanuel Beillard, Mélanie Gaillet, Céline Michaud, Véronique Servas, Nathalie Clement, Anaïs Perilhou, Thierry Carage, Didier Musso, Jean-François Carod, Stéphanie Eustache, Céline Tourbillon, Elodie Boizon, Samantha James, Félix Djossou, Henrik Salje, Simon Cauchemez, Dominique Rousset

**Author notes:** **Corresponding author:** Claude Flamand, Unité d’épidémiologie Institut Pasteur de la Guyane, 23 avenue Louis Pasteur, BP 6010, 97306 Cayenne Cedex, Mail. These authors contributed equally (Simon Cauchemez, Dominique Rousset).

## Abstract

**Background:** SARS-CoV-2 seroprevalence studies are crucial for clarifying dynamics in affected countries and determining the route that has already been achieved towards herd immunity. While Latin America has been heavily affected by the pandemic, only a few seroprevalence studies have been conducted there.

**Methods:** A cross-sectional survey was performed between 15 July 2020 and 23 July 2020 in 4 medical biology laboratories and 5 health centers of French Guiana, representing a period shortly after the epidemic peak. Samples were screened for the presence of anti-SARS-CoV-2 IgG directed against domain S1 of the SARS-CoV-2 spike protein using the anti-SARS-CoV-2 enzyme-linked immunosorbent assay (ELISA) from Euroimmun.

**Results:** The overall seroprevalence was 15.4% [9.3%-24.4%] among 480 participants, ranging from 4.0% to 25.5% across the different municipalities. The seroprevalence did not differ according to gender (p=0.19) or age (p=0.51). Among SARS-CoV-2 positive individuals, we found that 24.6% [11.5%-45.2%] reported symptoms consistent with COVID-19.

**Conclusions:** Our findings revealed high levels of infection across the territory but a low number of resulting deaths, which can be explained by young population structure.

## Introduction

The world’s attention remains focused on the spread of severe acute respiratory syndrome coronavirus 2 (SARS-CoV-2), that causes coronavirus disease 2019 (COVID-19), and the implementation of drastic control measures to limit its expansion. By the end of July 2020, more than 17 million confirmed cases and approximately 650,000 deaths have been reported worldwide [1]. With more than 4,500,000 cases and 190,000 deaths, Latin America has been particularly affected by the crisis [1].

A thorough evaluation of the proportion that has already been infected by SARS-CoV-2 and is likely immunized is important to estimate the level of herd immunity of the population [2] and to inform public health decision making. Data on laboratory molecular -confirmed cases do not capture the full extent of viral circulation because a majority of infected individuals have asymptomatic or mild infections and may therefore not seek care [3,4]. In contrast, population immunity is typically estimated through cross-sectional surveys of representative samples using serological tests. Numerous serological surveys conducted in affected countries at the beginning of the COVID-19 epidemic indicate that nationwide antibody prevalence varies between 1-10%, with peaks around 10-15% in heavily affected urban areas [5]. Most of the serological studies already available have been carried out in continental Europe [6–12] and in the United States [13–16]. Although Latin America has been heavily affected by the pandemic, only a few seroprevalence studies have been conducted across the continent, meaning the underlying level of infection remains largely unknown [5,17,18]. French Guiana, a French overseas department with 290,000 inhabitants [19], located in Latin America in the Amazonian forest complex experienced a large SARS-CoV-2 epidemic wave. A territory-wide lockdown was set up from March 17^th^ 2020 concomitantly with the rest of French territories, at a time when five imported cases and one secondary case were being confirmed on the territory [20]. The lockdown resulted in limited viral transmission until it was ended on May 11th 2020. In the middle of June there was a rapid intensification of viral circulation over a large part of the territory with 917 confirmed cases of COVID-19 detected from March 4^th^ 2020 to June 11^th^ 2020 [21]. This was followed by the implementation of strict mitigation measures such as curfews and local lockdowns in the course of June and July. The epidemic peaked at the beginning of July with 4,440 cumulative confirmed cases [22], followed by a gradual slowing down throughout the territory. Between March 4 and September 17, 9,623 cases (3,310 per 100,000 inhabitants) of COVID-19 and 65 hospital deaths (22.3 per 100,000 inhabitants) of COVID-19 were detected in French Guiana [23]. As the disease severity is reduced in younger individuals [24], we can suspect that many infections are likely to have been missed in this territory which has a much younger population (mean age of 25,1 versus 32,1 for Latin America and 42,3 for mainland France) [19]. In order to understand the underlying level of infection, we conducted a cross-sectional study within the general population, estimated the seroprevalence of SARS-CoV-2 and assessed its distribution in age groups and geographical areas.

## Methods

### Study area

French Guiana is composed of two main inhabited geographical regions: a central, urbanized area including a coastal strip along the Atlantic Ocean, where 90% of the population lives, and a more remote area along the Surinamese and Brazilian borders (Figure 1). This territory has the highest crude birth rate in the Americas (25.6 per 1,000 people) and 32% of the population is under the age of 15 [19]. Although living conditions are substantially more precarious than those of mainland France, French and European status gives French Guiana higher healthcare and diagnosis capacities than most South American countries. The healthcare system includes 8 medical biology laboratories and 3 hospital centers located in the main municipalities of the coastal area, as well as 17 healthcare centers located in more isolated areas.

**Figure 1.**
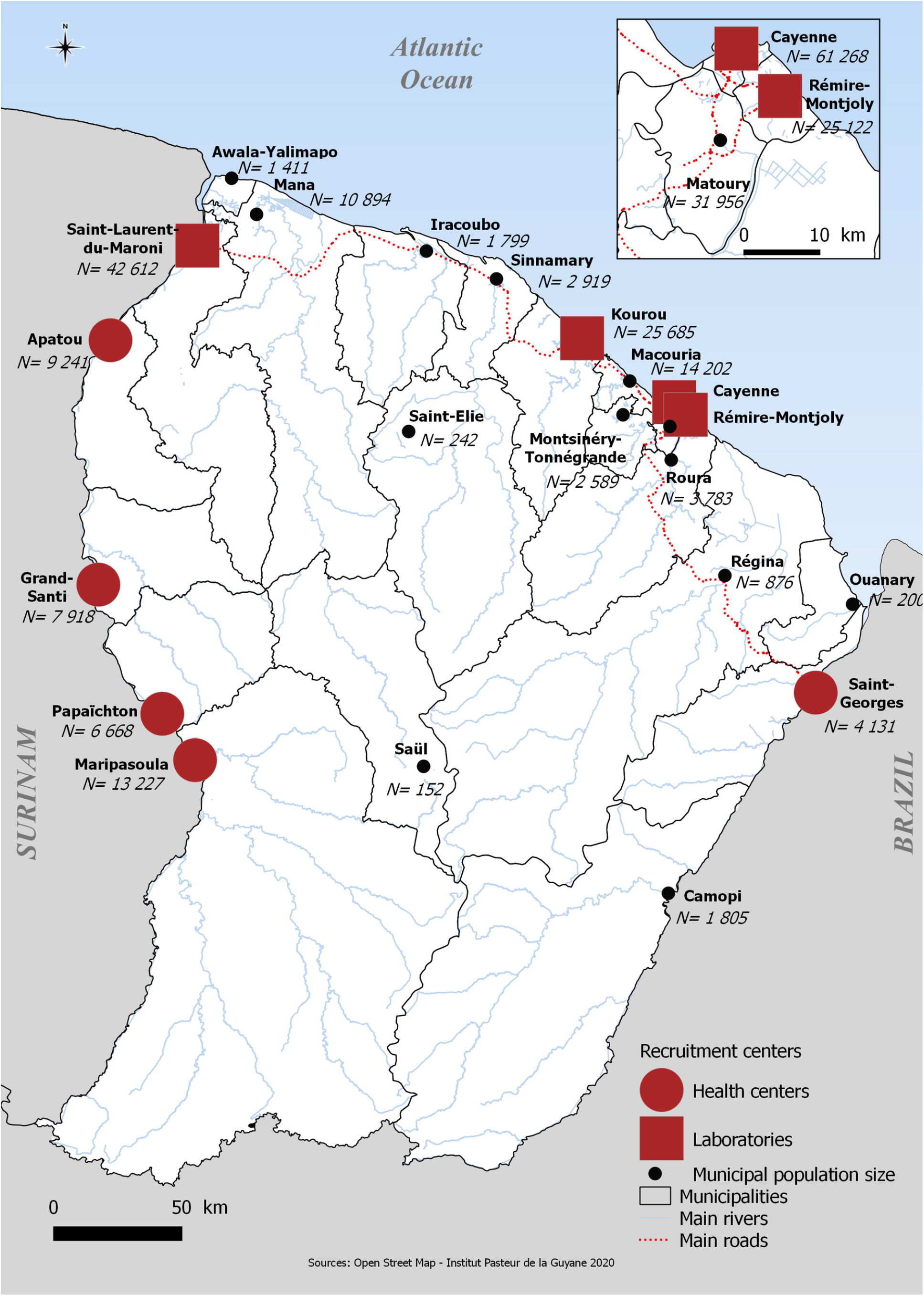
Map of French Guiana with municipal population size and recruitment centers.

### Study Design and Participants

A cross-sectional survey was performed between 15 July 2020 and 23 July 2020 in 4 medical biology laboratories located in the coastal urban area of French Guiana and in 5 health centers located in isolated areas along the Surinamese and Brazilian borders (Figure 1).

All individuals visiting the recruitment centers during the study period were invited to participate to the study, expected those admitted for SARS-CoV-2 RT-PCR testing.

### Procedures, ethical considerations

Publicity and information about the survey was provided at the reception desk of recruitment centers. Fieldworker investigators were trained to explain the project, collect informed consent and carry out the interviews. All individuals visiting the recruitment centers were invited to take part in the study during a preliminary face-to-face interview. For participants under 18 years-old, age-appropriate information was given and the consent of legally responsible person was collected. A specific educational-style comic poster was designed to explain, in an understandable way, the nature and objectives of the survey and inform them about the voluntary nature of the participation of the study.

Demographic data, including age, gender, residential region and occupation of each participant, were collected through a standardized questionnaire. Participants were asked to report the occurrence of a presumptive COVID-19 infection, to list the infection associated symptoms and to specify if they had consulted a doctor or obtained a biological confirmation of their infection. Thereafter, a venous blood sample of 3.5mL was collected from each participant, in accordance with current biosafety standards.

The study was recorded on Clinicaltrials.gov (ID: NCT04490850) and approved by the “CPP EST-III” Ethical Research Committee (No.CPP 20.07.04-8827; N°ID-RCB 2020-A01826-33). Personal data processing for this study comply with the requirements of the “reference methodology MR-001” established by the French Data Protection Authority (CNIL) regarding data processing in health research.

### Laboratory methods

#### Blood sample collection

Blood samples were collected into 3.5 ml gold BD Vacutainer SST II advance tubes with gel for serum separation (Becton-Dickinson, USA). Immediately after puncture, samples were stored at 4°C-8°C until centrifugation within 12 hours. Sera were then frozen and stored at - 20°C until used at the National Reference Center for respiratory viruses in Institut Pasteur in French Guiana.

#### Serologic diagnosis

Collected samples were screened for the presence of anti-SARS-CoV-2 IgG directed against domain S1 of the SARS-CoV-2 spike protein using the anti-SARS-CoV-2 enzyme-linked immunosorbent assay (ELISA) from Euroimmun (Lübeck, Germany). Semiquantitative results were calculated as the ratio of the extinction of samples over the extinction of a calibrator. According to the distributor, the specificity of the test was 99.6%. We internally validated the specificity of the serological assay with serum samples collected from 186 individuals of a cross-sectional serosurvey [25,26] in healthy individuals taken prior to the SARS-CoV-2 outbreak (June to October 2017). Of these, 40.8% were male and the median age was 33.3 years. In this validation subset, the serological test showed a specificity of 97.9% consistent of a recent assessment of this assay [27]. According to the distributor, the sensitivity of the test is 75.0% if the test is performed 10 to 20 days after infection and 93.8% if it is performed more than 20 days after infection.

### Statistical Analysis

In order to obtain population representative estimates of overall seropositivity in the territory we weight each sample by the population size within each municipality, age and sex group. We employ the following notation to describe the study design (Table 1):

**Table 1:** Weighted SARS-CoV-2 seroprevalence estimated by municipality, July 2020, French Guiana.

| Municipality (*i*) | *Pop.*<br>*size* | *Number of*<br>*enrolled*<br>*individuals* | *Weighted*<br>*seroprevalence*<br>*%[95% CI]* |
| --- | --- | --- | --- |
| Cayenne | 57,614 | 95 | 25.5% [10.9 – 48.8] |
| Matoury | 32,427 | 34 | 11.5% [3.5 – 32.2] |
| Saint-Laurent | 43,600 | 20 | 4.3% [1.0 – 16.8] |
| Kourou | 26,221 | 95 | 19.8% [8.6 – 39.1] |
| Remire-Montjoly | 23,976 | 98 | 4.0% [1.7 – 9.2] |
| Macouria | 11,719 | 10 | 20.7% [3.9 – 62.4] |
| Mana | 10,241 | 1 | 0 |
| Maripasoula | 11,856 | 13 | 15.2% [2.9 – 51.7] |
| Apatou | 8,431 | 62 | 12.1% [2.9 – 38.5] |
| Grand-Santi | 6,969 | 40 | 20.5% [8.7 – 41.1] |
| Saint-Georges | 4,020 | 29 | 16.1% [4.0 – 47.3] |
| Papaïchton | 7,266 | 10 | 7.8% [1.0 – 50.0] |
| Sinnamary | 2,957 | 2 | 0 |
| Roura | 3,713 | 8 | 0 |
| Montsinnery-Tonnegrande | 2,473 | 2 | 0 |
| Iracoubo | 1,878 | 1 | 0 |
| Total | 259,865 | 480 | 15.4% [9.3 – 24.4] |

- *i* : one of the 16 strata (municipalities);
- *M*_*i*_ : number of individuals living within the *i*^th^ stratum, *i=*1, …, 16 (Census data);
- *mi* : number of individuals enrolled from the *i*^th^ stratum, i=1, …, 16;

We considered that, in each municipality *i*, the probability of selecting a particular subject is equal to (1/ *m*_*i*_*/M*_*i*_) _*=*_ (*M*_*i*_*/m*_*i*_). This statistical weight indicates the number of people in the population represented by each subject in the sample.

We applied a post-stratification adjustment to each of these weights to arrive at the final statistical weight for each subject. This adjustment helped us to weight the age-sex groups within each municipality to match the distribution in the French Guiana total population. Nine age-groups ([0-15 years[, [15-20[, [20-25[, [20-25[, [25-35[, [35-45[, [45-55[, [55-65[, ≥65 years) were used within males and females groups and for each age-sex subgroups, we applied an adjustment factor *c*_*ijk*,_ to have a final statistical weight *w*_*ijk*_ *= (M*_*i*_*/m*_*i*_)^*\**^*c*_*ijk*_, where *i* indexes municipalities, j indexes sex groups and k indexes age groups. The outcome of interest was the weighted SARS-CoV-2 seroprevalence estimate. Analyses were carried out using survey capabilities of Stata version 15 statistical software [28]. Spatial distribution of seroprevalence was mapped using QGIS software [29].

## Results

We enrolled 480 participants between July 15 and July 23, 2020, in 16 municipalities (Table 1). The mean age of participants was 38.3 ranging from 0.2 to 87 years old. Comparison of the socio-demographic characteristics of the study sample to the census data demonstrated an over-representation of women (68.1% vs 50% in the general population of French Guiana) and adults over 25 years (79% vs 53% in French Guiana). These differences were corrected in the analyses of seroprevalence and risk factors by allocating a post-stratification weight to each participant.

Between July 15 and July 23 2020, the crude proportion seropositive was 13.2% and the overall weighted seroprevalence of SARS-CoV-2 antibodies in French Guiana was 15.4% [95% Confidence Interval CI, 9.3%-24.4%] (Table 1). This corresponds to 44,660 [95% CI, 26,970 - 70,760] seropositive individuals out of a population of 290,000.

Since the study was implemented two weeks after the epidemic peak and the sensitivity of the test is limited in the three weeks that follow infection, this likely represents a lower bound for the proportion of individuals infected by the time the epidemic peaked.

The seroprevalence did not differ according to gender (p=0.19) or age (p=0.51) (Table 2). Serological results in the different geographical areas (Figure 2) showed that SARS-CoV-2 circulated in most of French Guiana (Table 1). Highest infection risks were observed in the main population center in Cayenne municipality (25.5% [10.9% - 48.8%]) in the coastal and urban area. Two other municipalities located in the coastal area were also strongly impacted (Kourou: 19.8% [8.6% - 39.1%] and Macouria: 20.7% [4.0% - 62.4%]). However, even outside the main population centers, some areas appeared to have been heavily affected. For example, among the river areas, Grand-Santi (20.5% [8.7% - 41.1%]), a small municipality located in the western part of the territory and Saint-Georges (16.1% [4.0 – 47.3]), a municipality located in the eastern part, at the Brazilian border had seroprevalence levels similar to that observed in Cayenne (Table 2).

**Table 2:**
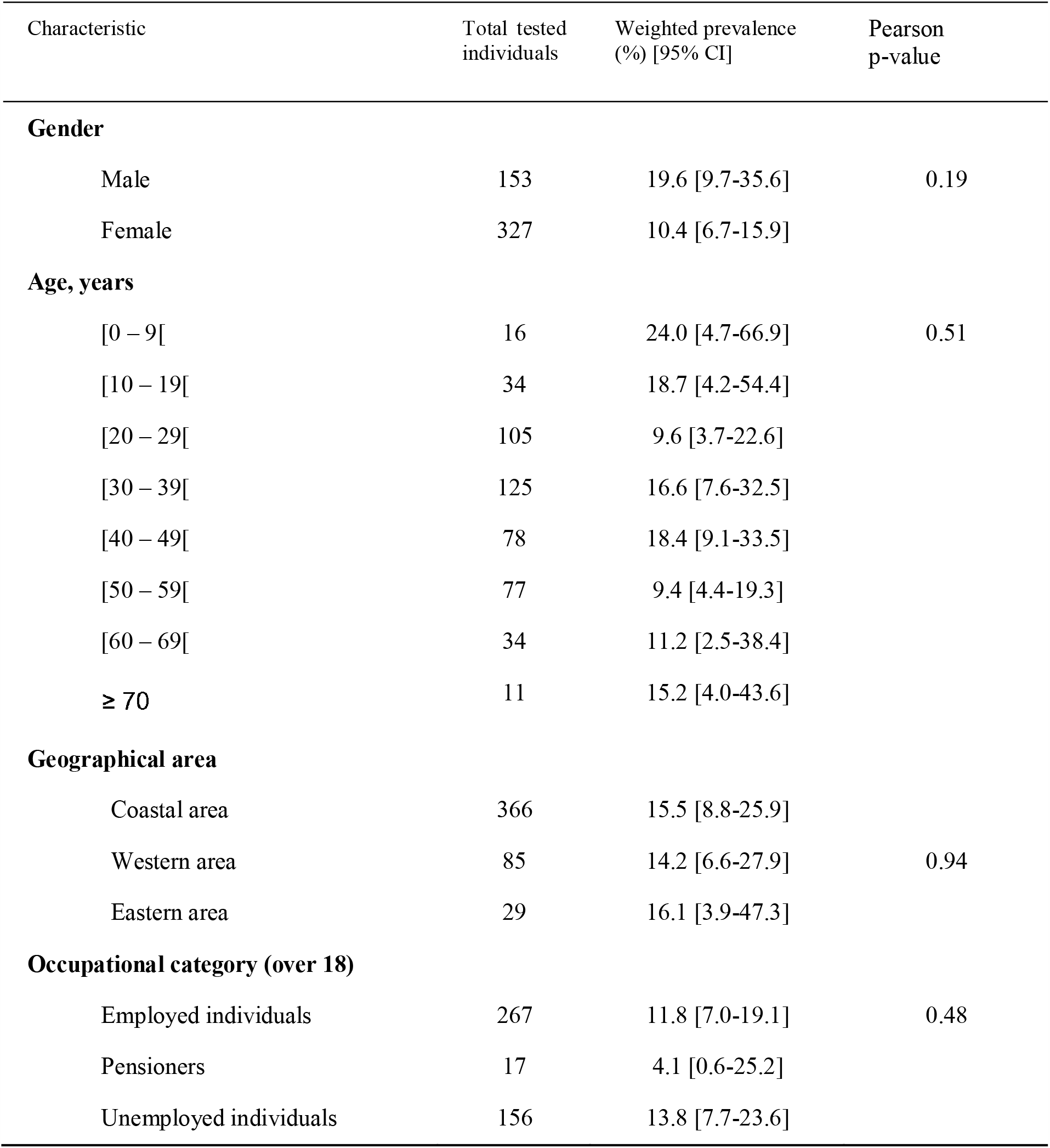
Distribution of SARS-CoV-2 seropositivity according to sociodemographic factors, July, French Guiana.

**Figure 2.**
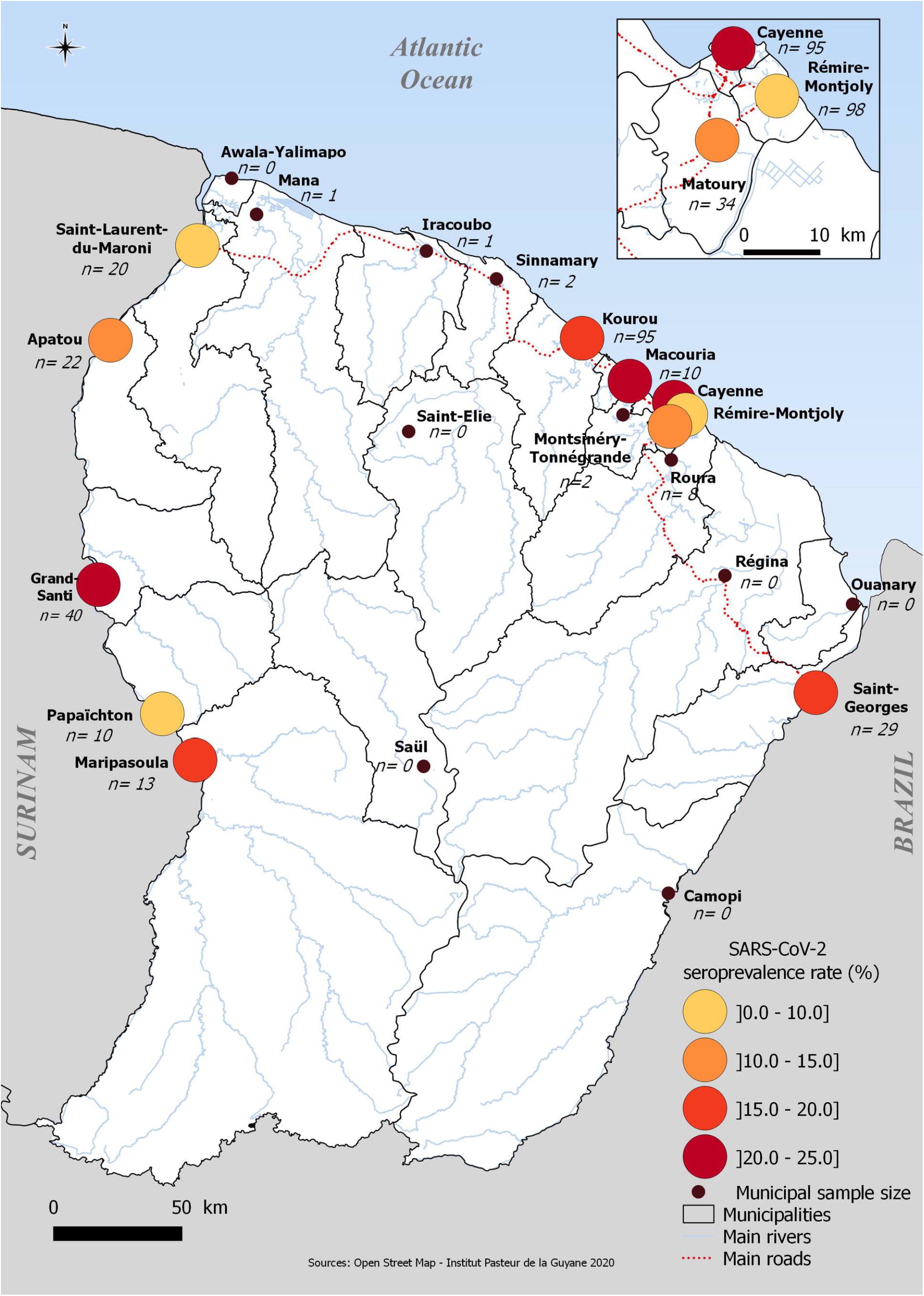
Spatial distribution of SARS-CoV-2 seroprevalence, July, French Guiana.

Thirty percent (29.9% [14.3%-52.3%]) of SARS-CoV-2 seropositive individuals reported they thought they had COVID-19 (vs 19.9% [14.4%-26.8%] of seronegative participants. Among those who reported a presumptive infection, twelve percent (12.6% [4.9%-28.7%]) declared that diagnosis was confirmed by a clinician and twenty percent (17.1% [6.9%-36.5%]) that the diagnostic was confirmed by a SARS-CoV-2 RT-PCR. Nine seronegative participants reported having a RT-PCR biological confirmation representing 2.1% [0.8%-4.9%] of seronegative participants. Among them, five reported infections less than 3 weeks old and three reported infections older than 3 months.

The presence of COVID-19 associated symptoms was reported in 24.6% [11.5%-45.2%] of seropositive participants vs 19.5% [14.0%-26.5%] of seronegative participants. Fever (50.7%), anosmia (48.8%), asthenia (48.8%) and cough (34.7%) were the most frequently reported symptoms in SARS-CoV-2 seropositive participants. However, only anosmia was significantly more prevalent in SARS-CoV-2 seropositive *versus* seronegative individuals (p=0.01) (Table 3).

**Table 3:**
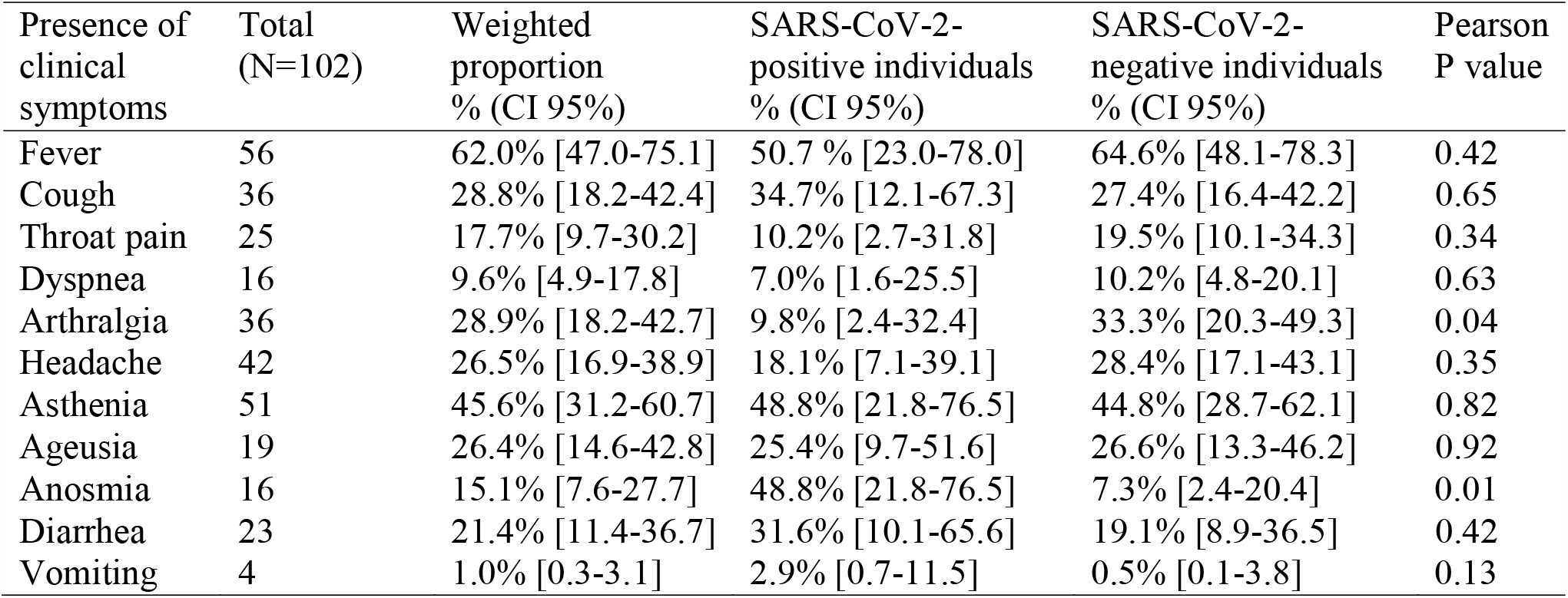
Clinical symptoms reported, by SARS-CoV-2 Infection status, July, French Guiana.

| Presence of clinical symptoms | Total (N=102) | Weighted proportion % (CI 95%) | SARS-CoV-2-positive individuals % (CI 95%) | SARS-CoV-2-negative individuals % (CI 95%) | Pearson P value |
| --- | --- | --- | --- | --- | --- |
| Fever | 56 | 62.0% [47.0-75.1] | 50.7 % [23.0-78.0] | 64.6% [48.1-78.3] | 0.42 |
| Cough | 36 | 28.8% [18.2-42.4] | 34.7% [12.1-67.3] | 27.4% [16.4-42.2] | 0.65 |
| Throat pain | 25 | 17.7% [9.7-30.2] | 10.2% [2.7-31.8] | 19.5% [10.1-34.3] | 0.34 |
| Dyspnea | 16 | 9.6% [4.9-17.8] | 7.0% [1.6-25.5] | 10.2% [4.8-20.1] | 0.63 |
| Arthralgia | 36 | 28.9% [18.2-42.7] | 9.8% [2.4-32.4] | 33.3% [20.3-49.3] | 0.04 |
| Headache | 42 | 26.5% [16.9-38.9] | 18.1% [7.1-39.1] | 28.4% [17.1-43.1] | 0.35 |
| Asthenia | 51 | 45.6% [31.2-60.7] | 48.8% [21.8-76.5] | 44.8% [28.7-62.1] | 0.82 |
| Ageusia | 19 | 26.4% [14.6-42.8] | 25.4% [9.7-51.6] | 26.6% [13.3-46.2] | 0.92 |
| Anosmia | 16 | 15.1% [7.6-27.7] | 48.8% [21.8-76.5] | 7.3% [2.4-20.4] | 0.01 |
| Diarrhea | 23 | 21.4% [11.4-36.7] | 31.6% [10.1-65.6] | 19.1% [8.9-36.5] | 0.42 |
| Vomiting | 4 | 1.0% [0.3-3.1] | 2.9% [0.7-11.5] | 0.5% [0.1-3.8] | 0.13 |

## Discussion

We report the first population-based serosurvey for the detection of SARS-CoV-2 antibodies in French Guiana. We found that 15.4% [9.3%-24.4%] of the population was seropositive two weeks after the peak of the epidemic. Assuming a two or three-week delay for seroconversion, our estimation reflects the level of infection of the population at the end of June or beginning of July, which roughly corresponds to the epidemic peak. Our results indicate that by that time, at least 44,660 [26,970 - 70,760] of French Guiana’s 290,000 population had been affected by the virus, more than 10 times the official count of 4,440 confirmed cases reported by public health surveillance system by the first week of July [22]. In Brazil, one of the bordering countries in French Guiana, seroprevalence estimates varied markedly across the country’s cities and regions, from below 1.0% in most cities in the south and center-west regions to up to 25.0% in the city of Breves in the Amazon (North) region [17]. Nevertheless, overall seroprevalence was estimated at 1.4% (95% CI 1.3-1.6). In contrast, our findings highlighted high but also relatively homogeneous levels of infection in most municipalities, ranging from 10% to 20%.

The case fatality rate of COVID-19 was low during the outbreak as there were 65 COVID-19 related deaths from the beginning of the outbreak up to September 17 [23] across the territory while about 45,000 people have been infected at the beginning of July. These was probably due in part to the young age of the population of French Guiana.

Younger populations are likely to have more social ties than older populations, and therefore physical distancing may be more difficult to implement than in countries with ageing populations. Furthermore, since young people are less exposed to disease severity, they may be less likely to adopt physical distancing measures when they are infected and potentially contagious in a context of high level of transmission.

Our study has several limitations inherent in the study design. Our approach made it possible to obtain rapid estimates of the impact of the epidemic. However, convenience sampling may result in a lack of population representation if part of the general population has lower access to the laboratories and health centers participating in the study. In our study, we observed a significant under-representation of men and children under 15 years of age, and therefore performed a post-stratification adjustment. However, this may have led to large confidence intervals for some of parameter estimates. In addition, sample size calculation was determined to obtain a sufficient point estimate of territory-wide prevalence estimates but not to study risk factors of infection. A few municipalities with no laboratory or health centers were not represented. However, the municipalities represented by the laboratories and health centers involved in the study are home to 80% of the population, so that our estimates are likely a good reflection of the situation across the territory. We may underestimate infection levels if precarious populations are at higher risk of infection and have limited access to health facilities.

Although specificity of the test used in our study is satisfactory according to our internal validation, we cannot rule out the possibility that a proportion of people infected at the beginning of the epidemic i.e. more than 3 months before our survey may have become seronegative [30]. In the other hand, it is possible that infected people did not develop specific SARS-CoV-2 antibodies or that these antibodies were not detected by our assay. Since the study was performed shortly after the peak of the epidemic, a proportion of individuals infected at the peak may not have seroconverted by the time of sample collection. With the exception of anosmia, symptoms were not significantly more frequently reported by seropositive than seronegative individuals. The high proportion of symptoms in SARS-CoV-2 seronegative participants may reflect the impact of a dengue epidemic that occurred concomitantly with the COVID epidemic in French Guiana [31].

In conclusion, we found that at least 15% of the population in French Guyana was infected by SARS-CoV-2 by the time the epidemic peaked in July. This corresponds to an elevated infection burden given the relatively limited mortality, which can be explained by French Guyana’s young population structure.

## Data Availability

Data are from the EPICOVID19 study belonging to Institut Pasteur, 25 rue du Docteur Roux 75724 Paris Cedex 15 France. Access to data is restricted for legal reasons according to the French CNIL recommendations (Commission Nationale Informatique et Libertes) that require specific authorizations to transfer health individual data from one center to another. The data may be made available after obtaining approval from the French regulatory authority: CNIL, Commission Nationale Informatique et Libertes, 3 Place de Fontenoy TSA 80715 75334 PARIS CEDEX 07, France. Phone (33): 01 53 73 22 00. Request from data transfer can be sent to Clinical Core of the Center for Translational Science of Institut Pasteur, Paris, Tel : + 33 (0) 1.40.61.38.74; Fax : + 33 (0) 1.40.61.39.77; https://research.pasteur.fr/en/team/clinical-core/

## Acknowledgments

We are grateful to all field workers, collaborators, technical and medical staff from biological laboratories and health centers involved in the EPI-COVID-19 project. We thank Bhety LABEAU, David MOUA, Laetitia BREMAND, Elisabeth CHAN from Institut Pasteur in French Guiana, Nathalie JOLLY from Clinical Core of the Center for Translational Research of Institut Pasteur. We also thank Clara pichard, Veronique, Céline, Roxane Schaub, Sylvain Fradin, Beatrice Pesna, Anna Edwige, Fabien Rogalle, Jean Yves Cattin, Nicolas Lormee, Véronique Olin, Charlène Cochet, Clara Fernandes from Health Centers Department of Cayenne Hospital Center.

